# STILLBIRTHS: PREVALENCE, RISKS AND CLINICAL INTERVENTIONS IMPACT AMONG WOMEN IN KWAHU GOVERNMENT HOSPITAL

**DOI:** 10.1101/2025.02.18.25322461

**Authors:** Gaddafi Abdul-Salaam, Michael Owusu-Adjei, Kwakye Ansong, Joseph Manasseh Opong, Nana Osei, Ampadu Seth

**Affiliations:** Department of Computer Science, Kwame Nkrumah University of Science and Technology, Kumasi, Ghana; Department of ICT & Mathematics, Presbyterian University, Ghana; Department of Obstetrics and Gynaecology, Kwahu Government Hospital, Atibie, Ghana; Departments of ICT & Mathematics, Presbyterian University, Ghana

## Abstract

**Background:** It is the expectation and joy of every expectant mother and her immediate family to welcome into their fold healthy newborn babies after each gestational period. Unfortunately, in most instances this expectation is marred by complications which ocassionally result in the death of the unborn/newborn child known as stillbirth. This anti-climax leads to unexpected periods of sadness for both mother and her immediate family members. The growing incidence of stillbirths is acknowledged worldwide and said to be associated with many factors both maternal and child health conditions. However, the key question is how many of these maternal and child conditions could have been anticipated and prevented. Addressing this challenge will require a review of specific instutional policy frameworks that deals with the incidence of stillbirths on a case by case basis. In this retrospective study, record of stillbirths over a 5-year period is examined to determine impact of clinical interventions on prevalence and risks among women within Kwahu West municipality in Ghana.

**Method:** A retrospective cross-sectional study to examine recorded cases of stillbirths over a 5-year period among women in Kwahu South Hospital to determine clinical intervention impact on prevalence, risks and control was undertaken. Using established clinical audit recommendations, this study examines the effectiveness of recommendations and its impact. Socio-demographic features such as age, educational level, gestational maturity, medical conditions of mother and child, delivery method, recorded stillbirth types, fetal sex and patient’s geographical location. All statistical analysis was performed using R programming tool.

**Results:** Clinical interventions and its impact show a reduction in stillbirth types from 4 to 2 from 2019-2023. However, macerated and fresh stillbirths remain the most dominant stillbirth types recorded with and without medical conditions in urban communities. Sustained decreases in the number of stillbirths across all years including the elimination of early neonatal deaths and intra-uterine fetal deaths from 2021-2023 is observed.

**Conclusion:** Established institutional clinical audit on each stillbirth occurrence provided in-depth analysis on case-by-case basis. This practice led to the use of clinically designed interventions resulting from audit reviews on specific cases to help address similar situations. Even though a reduction in the number of recorded stillbirths from 20.83 per 1000 live births in 2019 to 10.81 per 1000 live births in 2023 is noted, a staggering 76.88% share of stillbirths among patients and fetus with no medical condition presents a serious challenge to healthcare providers. Detailed review of all cases that excludes nothing irrespective of clinical presentation can provide important insight into the incidence of high stillbirth rates among women with such presentation.

## Introduction

There is growing concern for increasing deaths at or after 28 weeks of pregnancy. This concern is evident in the number of recorded at or after 28 weeks of pregnancy. It is estimated that [1] in 2021, the total number of such deaths recorded reached 1.9 million but many of which could have been prevented with adequate or proper care. Globally, the death rate of 13.9 per 1,000 births exist for such situations but geographical inequalities make it more or less prevalent in different societies than others [2]. The risk of death at or after 28 weeks of pregnancy in sub-Saharan Africa as shown by the growing number of recorded cases. There was 0.77 million deaths in 2000 as against 0.82 million in 2019 [3]. It is estimated that, stillbirth rates (high or low) is a reflection of the quality of care given (antenatal care, timeliness of care and quality of intrapartum monitoring and care provided) [4]. This reflection of quality of care is highlighted in a study, which assessed the impact of improving antenatal care. Study conclusions show that, instituting preventive measures and promoting deliveries in healthcare institutions where anticipated complications can be better managed has the potential of reducing stillbirths by almost half [5].

Experiences among women and families who suffer stillbirth effects vary from one socio-cultural context to another but its involvement in psycho-social morbidity is emphasized [6]. Different causes of stillbirth which include maternal and child conditions such as pre-eclampsia, antepartum haemorrhage, placenta abruptio, fetal anomaly, abnormal fetal presentation, uterine rupture, fetal distress, arm presentation, intra-uterine growth retardation, prolong labor etc has been emphasized in many research works. Such studies include a qualitative research to determine associative factors with stillbirth which listed among other factors, available and accessible medical care facilities, conduct of healthcare givers during pregnancy, in labor and after delivery care services as important factors to help reduce the incidence of stillbirth [7]. In its assessment of coverage areas of concern, patient’s standard of care [8], quality of care given to mothers and newborn infants, proportion of stillbirths which met pre-defined standard of care and could have been saved and recommendations for improvements in stillbirth audit process constituted legitimate areas of concern for investigations to highlight important contributory factors. Further case study approach to identify incidence and determinants of stillbirth highlighted prevalence rate of 31.3% per 1,000 births within the stuy area. Pre-eclampsia was the commonest medical condition for the incidence of stillbirth among the study population [9]. The changing nature of disease symptoms and disease diagnosis shows that adopting institutional protocols with detailed procedures to help deal with the incidence of stillbirths on a case-by-case basis will help address rising stillbirth rates. This research takes a 5-year record of stillbirths, examines institutional protocols detailing procedures for addressing incidence of stillbirths and shows how this approach has influenced prevalence, risks and controlled stillbirths over a 5-year period.

### Related research works

Hightened interests in stillbirth as a serious public health concern has had extensive research investigation many of which is aimed at estimating risks factors and determining prevalence rates among sections of population. This section explores many of these related works to determine research gaps. From its estimation of stillbirth rates from observational studies, an estimated incidence rate of 180 stillbirths per 1,000 deliveries among women in Kano, Nigeria [10]. Risks identified included lack of maternal education, history of previous stillbirth(s), prematurity, socio-demographic factors such as living in both semi-rural and rural settings and having extended periods between rupture of membranes and delivery. Additional findings in a multivariate analysis wihin the same study show further excerbation of the problem with women who had no education and showed no sign of disease. Increasingly high incidence of stillbirths as amplified by additional studies show 175 stillbirths from 4416 deliveries in referral hospitals in Nigeria. It estimated that 22.3% of the recorded stillbirths was macerated, 47.4% constituted fresh stillbirths and 30.3% were unclassified and attributed the high incidence to patient related factors [11]. An estimated 30.7 per 1,000 stillbirths because of low maternal education, history of previous stillbirth, pre-term delivery and incidence of child congenital anomalies as mentioned in a cross-sectional study in Ethiopia [12]. Similarly, high maternal age, low maternal education, and rural habitation were factors determined to be associated with stillbirths among women sampled [13]. A related cross-sectional study to determine stillbirth risk factors among women with severe pre-eclampsia found prevalence rate of 9.8% from a study population of 469 women [14]. Lack of knowledge, cultural practices, and attitudes of health workers are perceived to be contributory factors for the cause of stillbirth in a qualitative study that explored community perceptions of stillbirth [15]. In its assessment of impact from maternal medical diseases and obstetric complications using retrospective study design approach [16], prevalence rate of 6.2 per 1000 live and stillbirths was determined. Risks of increase in stillbirth is attributed to maternal medical conditions such as diabetes mellitus, chronic hypertension, pregnancey complications such as intrauterine growth retardation/restriction and amniotic fluid index measurement levels (oligohydromnios/polyhydromnios). Features listed among key indicator points for stillbirth was fetal growth restriction. Advocate for antenatal surveillance and early delivery to reduce the risk of stillbirth in pregnancies with suspected or confirmed fetal growth restriction is recommended [17] in a related research study which called for the evaluation of fetal growth restriction with fetal autopsy, placental pathology, genetic testing, antiphospholipid antibody testing, and fetal-maternal hemorrhage testing. Additional features of stillbirth is determined from a study that identified education level, marriage age, age of first conception, number of children, consanguineous marriage, employment status, and diseases like diabetes, hypertension, and history of depression as significant predictors [18]. A prospective, population-based study involving many countries also determined high risk of stillbirths [19] among women found to be less educated and less antenatal/obstetric care patients going for vaginal assisted deliveries. The impact of improving quality of antenatal care and its effect on stillbirths as listed among specific maternal healthcare interventions to reduce the risk of stillbirths include antenal care attendance; targeted aspirin prophylaxis, ideal sleep positions, fetal movement monitoring, preconception and interconception care etc [20].

Disparities exist in stillbirth rate as recorded in a related study that used 2,868,482 singleton births, to determine overall stillbirth risk. A stillbirth rate of 3.1 per 1000 live births is recorded among women with increasing body mass index in early and after 39 weeks of gestation [21]. Similarly, in its review of primary records of 168 child deaths, decisions made about diagnosis and management in primary care were determined to affect the cause, time and circumstances of a child’s death [22]. Based on the combination of current pregnancy complications, congenital anomalies/malformations, maternal characteristics, and medical history, stillbirths can be identified at the level of antenatal care [23].

Research into the incidence of stillbirth is fraugth with both financial costs and recruitment problems. 169 out of an expected 254 eligible patients participated in a study to assess factors influencing recruitment in a multicentre case-control stillbirth study [24][25]. Despite, these challenges, efforts continue to be made for research into stillbirths to provide the needed information regarding incidence, risks of current and future pregnancies and its management, family planning measures, delivery process management, effect and impact of clinical care interventions on pregnancy related disease control practices and many others that can bring about better management of child delivery.

### Data Analysis

R language is an open-source programming tool ideal for statistical data visualization. The tool used for all data analysis and data visualizations is the R-programming tool. Data visualizations, which include graphs, boxplots, scatter plots, histograms, provides detailed information about the incidence of stillbirths extensively discussed in the results section. Exploratory data analysis with results presented in plots of graphs, frequency count distributions, histograms etc.

### Definition of Features

Level of education (lev_edu) in this research: Is determined to be the patient’s highest point of educational attainment. (h_sch is senior high school, j_high is junior high school, primary is basic school, none means no formal education, tertiary includes all post senior high school qualifications including professional certifications).

Medical condition (med_condition): Is determined to be inclusive of both fetal and maternal. This includes pre-medical conditions of maternal.

Location: Is defined as the maternal town/city of habitation

Fetal sex (f_sex): determined as the sex of the stillborn

Maturity in weeks (maturity_wks): Maternal Maturity in weeks

Stillbirth status (stl_status): Conditional classification of the stillborn

Mode of delivery (m_del): Maternal mode of delivery

Year: represents the recorded year of occurrence

Age: maternal age in years

## Materials and Methods

This retrospective case study approach considered recorded incidence of stillbirths over a 5-year period spanning from 2019-2023. Some of the exclusion criteria were records of stillbirths and data on total number of transfer-in patients from the various healthcare clinics, private health institutions and other centers within the smaller distant communities in the municipality. Data collected included in-patients, outpatients and transfer in patients. The study also examined detailed record of clinical audits including specific recommendations on a case-by-case basis. Kwahu Government hospital is a fully funded government of Ghana institution initially established by the Seventh Day Adventist Church in the 1950’s. The largest referral healthcare institution which serves an estimated population of over 200,000 consisting of diverse ethnicities, professional and religious orientations but predominantly traders, fisher folks and farmers. This healthcare institution boost of two well experienced clinical Obstetric and Gynaecological specialits whose work experience span over 15 years at this facility. It has modern equipments such as advanced ultrasound machines of various types, cardiotograph machines (CTG), facilities to manage pre-term infants which includes incubators. Easy to reach access to its location makes it an ideal referral point of call for all types of referral emergencies. The institution offers important healthcare services such as antenatal clinics, postnatal clinics, elective surgeries, radiology services including daily and on-demand ultrasound services of all types such as fetal anomaly scans and many others.

The Department of Obstetrics and Gynaecology in collaboration with the Antenatal clinic over the last 10 years has institutionalized and operationalized “club 36 School”. This ‘school’ focuses on educational programs which include; nutritional requirements for pregnant mothers, education on signs and symptoms of labor, birth preparadness and complications readiness, familiarization tour with facilities at the labor ward, operating theatre, maternity wards etc, for expectant mothers who reach 36 weeks maturity for both nulliparous and multiparous women.

### Ethical Approval Considerations

Approval for the conduct of this research is based on rules and guidelines set out in the ethics review committee on research involving human subjects’ policy document by Ghana Health Service. Request for approval to conduct and use retrospective data of recorded stillbirths and minutes of records from the clinical audit committee for the period january-december (2019-2023) was granted with approval number KGH/GHS/30/24. This request excluded patient names and other identifiable features necessary to safeguard the privacy of patients. Assessing the impact of interventions strictly used recommended data as contained in the audit committee reports, which included specific recommendations for future case management.

## Results

Summary of descriptive analysis showing record of stillbirths grouped by years and the cummulative record of stillbirths combined for the study period of 2019-2023 in Table_1_2 with 173 cases reported during the period of 2019-2023. In 2019, 42 cases made up of fresh and macerated stillbirths, 46 cases in 2020 made up of fresh, macerated intra-uterine deaths and early neonatal stillbirths, 44 cases in 2021 made up of fresh and macerated stillbirths, 18 cases in 2022 consisting of fresh and macerated stillbirths and 23 cases in 2023 with fresh and macerated stillbirths. Summary statistics is of total stillbirths is reported in Table_1_2 which shows frequency counts stillbirth types and percentages obtained by each count type. Combined graphical display of stillbirth types recorded under each reviewed year is shown in Fig 1. With the exception of year 2020, which show four stillbirth types (fresh, macerated, intra-uterine and early neonatal stillbirths), only two stillbirth types (fresh and macerated stillbirths) were recorded for the year 2019, 2021, 2022 and 2023 respectively. Maternal age graphs in Fig 2 show the distribution of age and Fig 3 represents scatter plot of maternal age distribution. Recorded minimum and maximum ages were 12years and 47 years with a standard deviation of 6.89. Distritution of frequency counts of stillbirth types over the study period from 2019-2023 is shown in Fig 4. Highest level of educational attainment is shown by a distribution graph as image_1. This graphical distribution show patients whose formal education started from primary to the tertiary level. Educational Levels completed are assigned as primary, junior high school, senior high school, tertiary and those who have never had formal education as none. Recorded reasons in the form of medical conditions and the frequency count of occurrence of stillbirths as shown below is also described in a frequency count graph in image_2.

**Fig 1.**
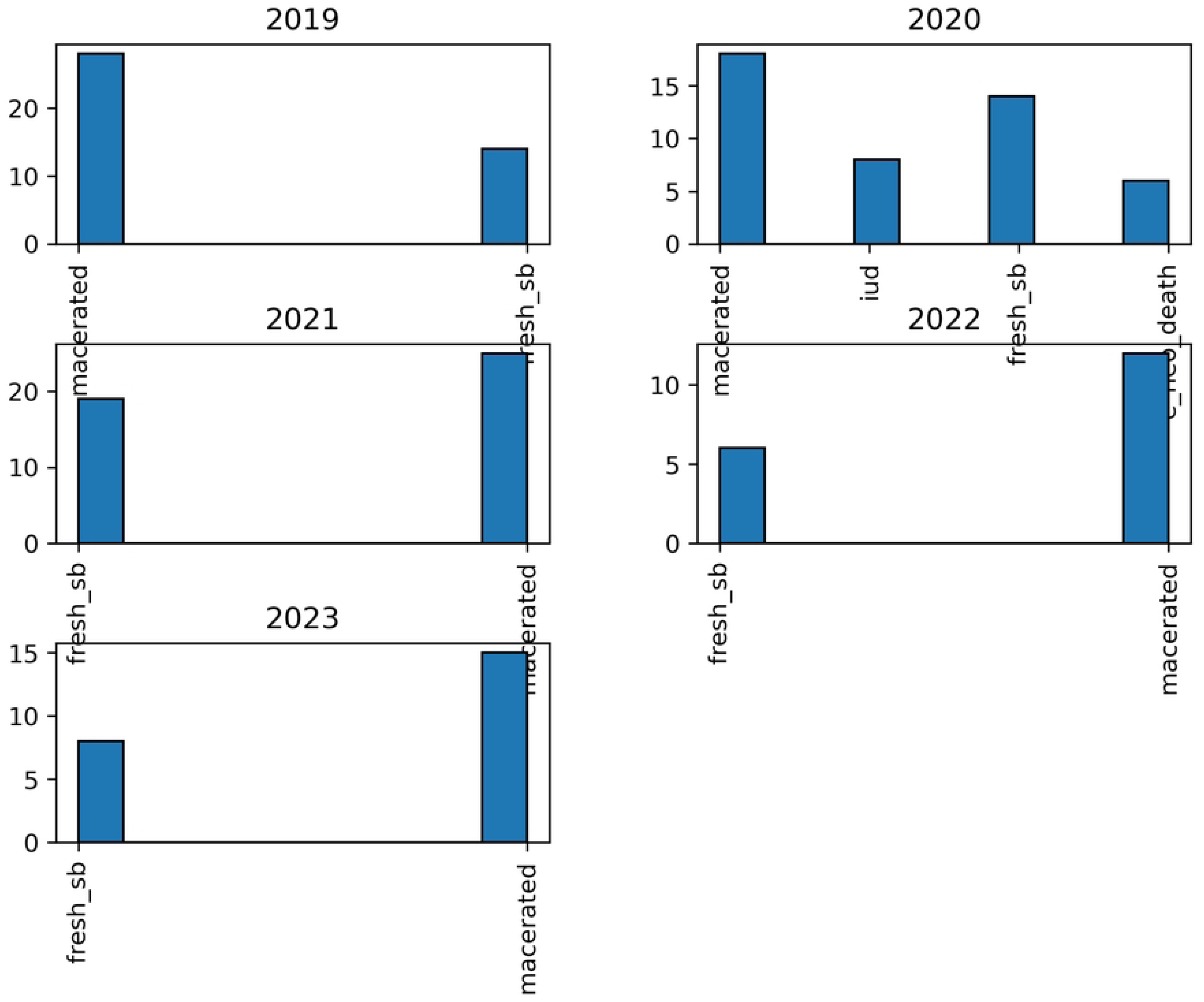
Stillbirth type graphs. **Caption**: Individual graphs show stillbirth types over the 5-year period

**Fig 2.**
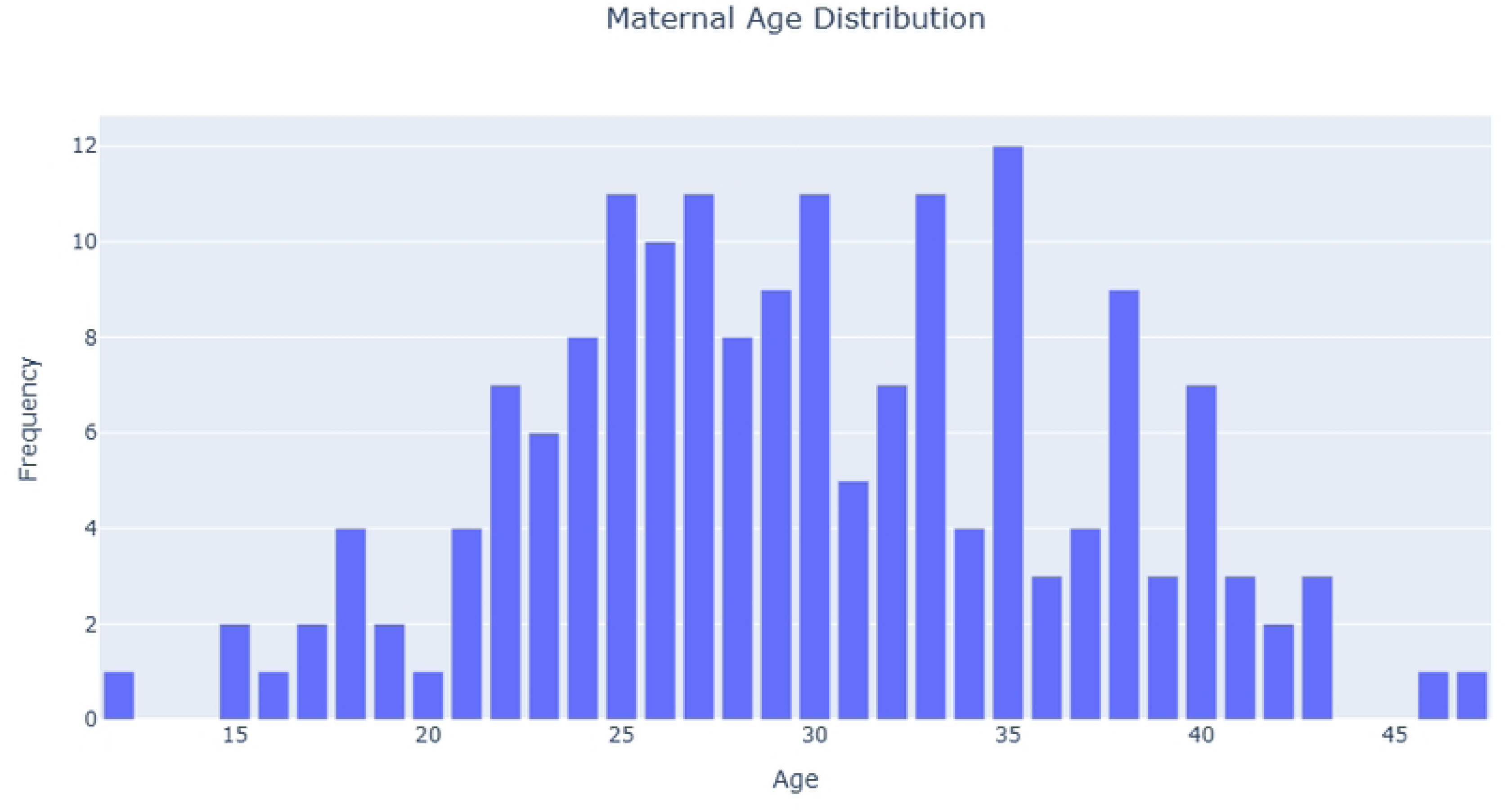
Maternal age distribution graph. **Caption**: Maternal age distribution of the study sample

**Fig 3.**
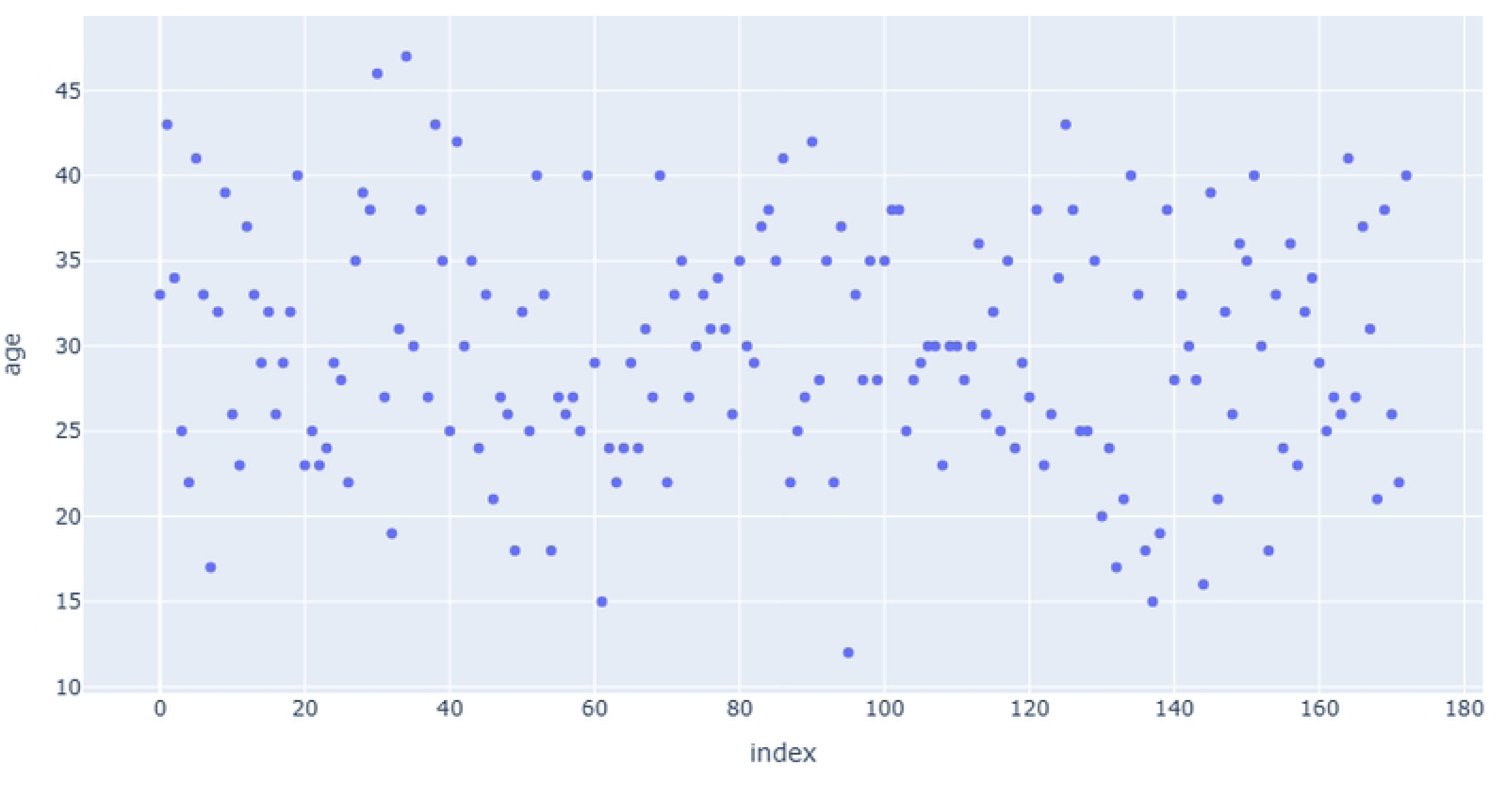
Maternal age distribution scatter plot. **Caption**: Presentation of maternal age distribution in a scatter plot.

**Fig 4.**
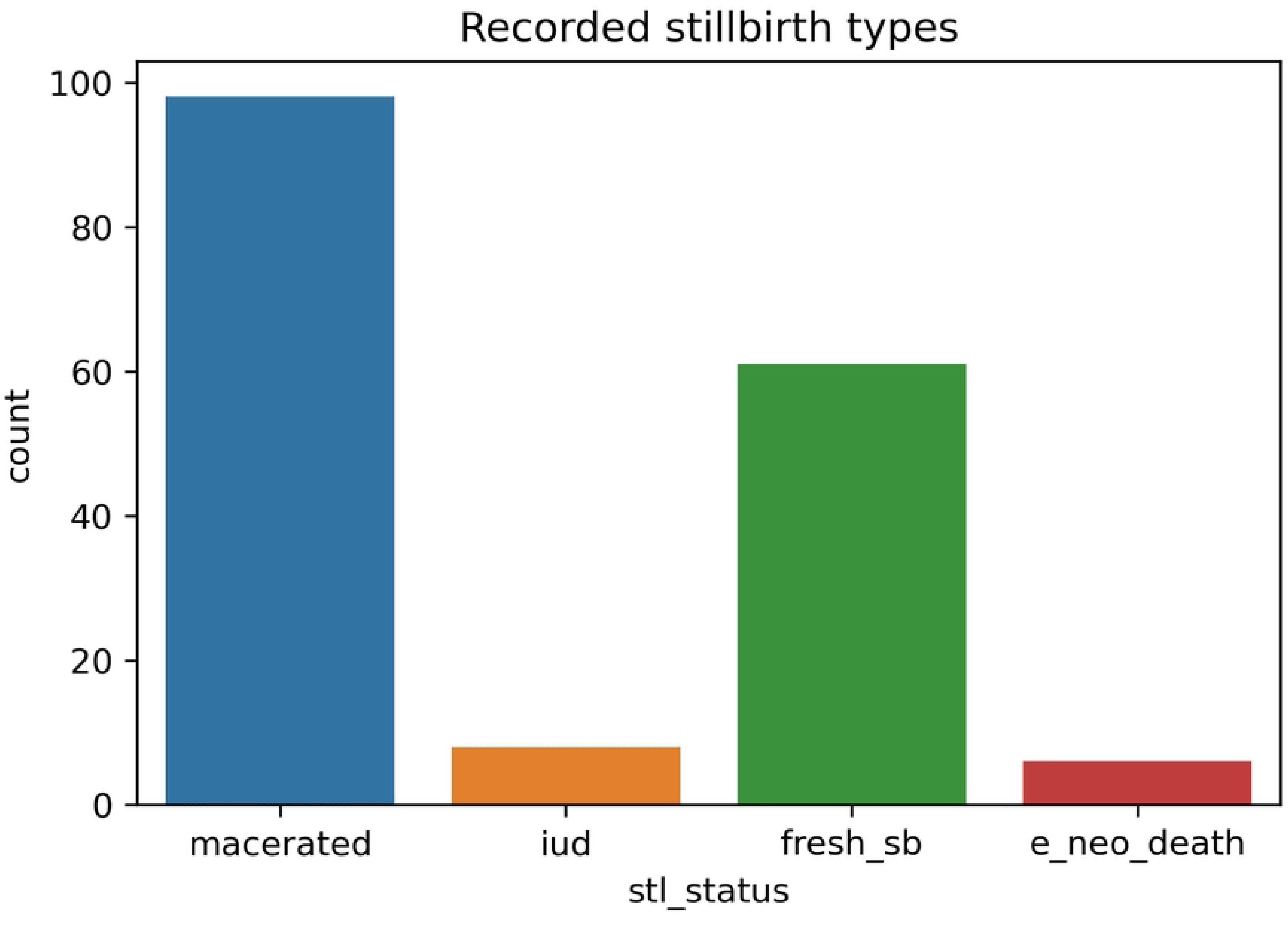
Combined stillbirth types: **Caption**. Stillbirth type combined graph to demonstrate the count of each stillbirth type.

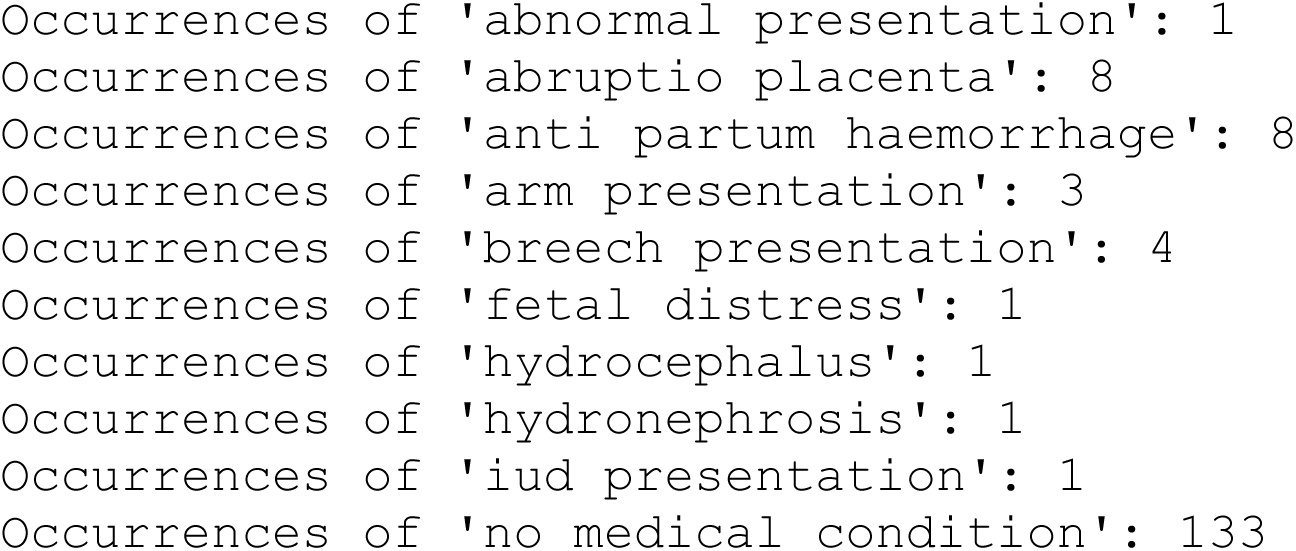
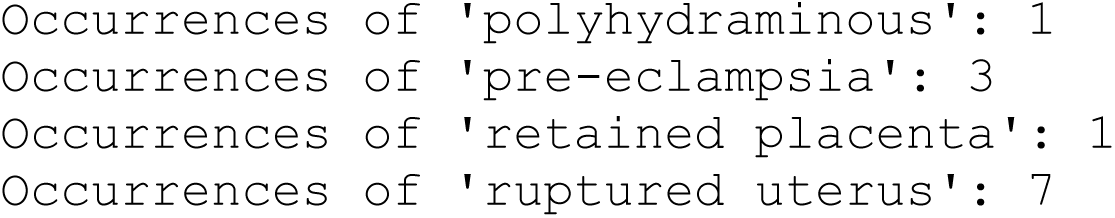

### Prevalence of Stillbirths

A total number of 173 stillbirths were recorded over the study period from 2019-2023. Birth deliveries over the same period were 2,016 for 2019, 2,215 for 2020, 2,215 for 2021, 2,102 for 2022 and 2,128 for 2023. Rate of stillbirths per 1,000 live births were; 20.83 for 2019, 20.77 for 2020, 19.86 for 2021, 8.56 in 2022 and 10.81 in 2023. 133 stillbirths, constituting 76.88% from the total number of 173 stillbirths occurred with no record of medical conditions for either mother or child. Incidence of stillbirth and prevalent maturity in weeks of occurrence is shown by image_3 boxplot. Ratio of proportion on educational level of attainment on stillbirths as shown in image_4, defines a relationship between stillbirth types and maternal level of education. Prevalent rates show that early neonatal deaths is associated with women whose educational levels ranges from primary, junior high and those without formal education marked as none. However, intra-uterine fetal deaths were closely associated with women whose educational levels were junior high school and tertiary.

It is noted that, recorded incidence of stillbirths for the study period show no occurrence below or at 32 weeks of gestation Maternal geographical location in the study area and its relationship with the incidence of stillbirths is recorded in image_6. High incidence of macereted and fresh stillbirths among women in urban communities is shown by the high frequency counts in image_6.

In the trend analysis of recorded stillbirths by year, image_6 shows the impact of clinical decision interventions. Apart from year 2020, there have been no reported cases of early neonatal and intra-uterine fetal deaths including declining levels of macerated and fresh stillbirths as shown in the distribution of stillbirths by year image_7 from 2022-2023.

### Risk factors for Stillbirths

Even though the incidence of stillbirths, continue to decline since 2021, challenges remain for addressing risk factors. In the bivariate analysis, high incidence of stillbirths among women with no history or evidence of maternal or child medical condition is recorded. In image_2, distribution of stillbirth by medical condition show high incidence among women who showed no sign of adverse medical or child condition. Advancing maturity (in weeks) above 32^+^ makes it an important risk factor as demonstrated in image_3 boxplot and this is supported by the gestational maturity of clinical audit records of stillbirths in Table 3. Additionally, high incidence in densely populated locations as shown in image_5 makes socio-demographics an important risk factor to consider contrary to study findings [9] which established no relationships between patient’s socio-demographics and the incidence of stillbirth.

**Table 3:** Clinical audit and interventions

| Patient condition | Indication | Maturity/weeks | Outcome | Recommendations/Interventions |
| --- | --- | --- | --- | --- |
| Postdate | Slow progress of labor | 43 | Fresh sb, CS | Intervention by clinical staff<br>(midwife/gynaecologist) |
| Known Hypertensive | G3P2A | 37 | Macerated, SVD | Early delivery to avoid complications |
| Lower Abdominal Pain | Non-assuring fetal heart rate | 37 | Fresh sb, CS | CTG, interpretation |
| Nullip | Losing liquor | 44 | Macerated<br>SVD | Recommend early delivery to avoid placenta calssification. |
| Breech presentation | FHR 134 bpm | 37 | Fresh sb, CS | Immediate attention to avoid prolong labor and uterine rupture |
| Previous salpingectomy | None | 35 | Early neonatal,<br>SVD | CTG monitoring |
| Symphysial Fundal Height, 38cm | No FHR | 39 | IUD | Screen for Chronic diseases |
| Symphysial Fundal Height, 42cm | Breech presentation | 39 | Fresh sb, SVD | Regular Blood Pressure check-up, avoid SVD for high SFH patients. |

In Table 3, presentation of patient characteristics before, during and onset of labor provides evidence-based clinical decision support interventions to determine the course of action on case scenarios.

## Discussions

In the 5-year trend analysis of incidence, declining stillbirth rates from 20.83 in 2019 to 10.81 in 2023 per 1000 live births [26] demonstrates strong impact of clinical decision interventions arising from clinical review audit recommendations of all stillbirths at the designated healthcare facility. This and many other research works have demonstrated several causes of stillbirths some of which include, pre-eclampsia, fetal distress, congenital malformations, antepartum haemorrhage, advanced maturity in weeks of above 40 and many others. However, challenges with increasing incidence of stillbirths among women with no known adverse medical condition [27][28] remains a challenge. Of a total 173 stillbirth reviewed in this study, 133 constituting 76.88% is attributed to women/fetus with no known medical/clinical condition. Contrary to reported effect and associations of formal education on stillbirth rates as reported[29][30][31], analysis carried out in this study show no correlation between formal educational attainment and the incidence of stillbirths. However, incidence of certain types of stillbirths is reported among women in particular educational levels which include no formal education [32]. The high incidence of stillbirths among women with high maturity [33], no known adverse medical condition and in urban geographical locations is under-scored in this research study.

### Study Strengths and Limitations

This study has demonstrated how clinical audit recommendation implementations have affected the incidence of stillbirths over a 5-year study period. It has also brought to the fore, the threat and increasing incidence of stillbirth among women/fetus with no known medical condition. By including formal educational levels attained by each patient, the study has shown that increasing/reducing stillbirth incidence is not associated with one’s level of education but rather, certain stillbirth types could be associated with certain educational levels. It is noted in this study that, being located in a rural community puts no patient at a disadvantage as increasingly high number of stillbirth incidence analysed in this study had happened in women in urban communities. Limitations in this study include non-inclusion of fetal weight, maternal body mass index, information on total number of transfer-in patients from other healthcare institutions and healthcare centres in the rural communities and urban centers.

## Conclusions

Analysis made and conclusions drawn in this study are based on empirical data collected at the designated wards and departments which were the focus of audit presentations at the stillbirth clinical audit meetings. Among a list of medical conditions examined to be associated with the incidence of stillbirths in this study, higher incidence of macerated stillbirths is determined to be associated with mothers with no medical conditions. This concern is shared in other research studies as well [34][35]. Analysis of incidence per maturity shows the incidence of stillbirths above 32 weeks of maturity as indicated in image_3

Contrary to other research findings [29], analysis undertaken in this study found no significant disparities between mothers educational level and the incidence of stillbirth as demonstrated in image_4. However, disparities in macerated stillbirths exist between women in urban communities and those in the rural areas as shown in image_5. The ratio of proportion of stillbirths over the period shows a drastic reduction in recorded intra-uterine deaths and early neonatal deaths. However, challenges exist for the incidence of macerated and fresh stillbirths especially among women with no medical conditions. This phenomenon and the incidence above 32 weeks of maturity, therefore calls for a deeper assessment of all pregnancies to identify potential risk factors from among women who presents no medical condition.

## Declarations

**Authors Contributions**: Conceptualization, Michael Owusu-Adjei,

Methodology: Gaddafi Abdul-Salaam, Oppong Manasseh

Supervision: Nana Osei, Kwakye Ansong

**Funding**: This publication is not funded by any organization or individual.

**Ethics approval and consent to participate;** Approved

**Consent for publication:** Approved

**Availability of data and Materials**: Dataset used for analysis is available through (https://github.com/owusuadjeim/owusuadjeim/blob/main/stbth_data.csv).

**Conflicts of Interest**: The author(s) declare that there is no conflict of interest regarding the publication of this paper.

## Data Availability

Data is available at https://github.com/owusuadjeim/owusuadjeim/blob/main/stbth_data.csv

## Acknowledgments

Appreciation goes to the management and staff of Kwahu Government Hospital for their immense support and assistance.

---

**Image_1.**
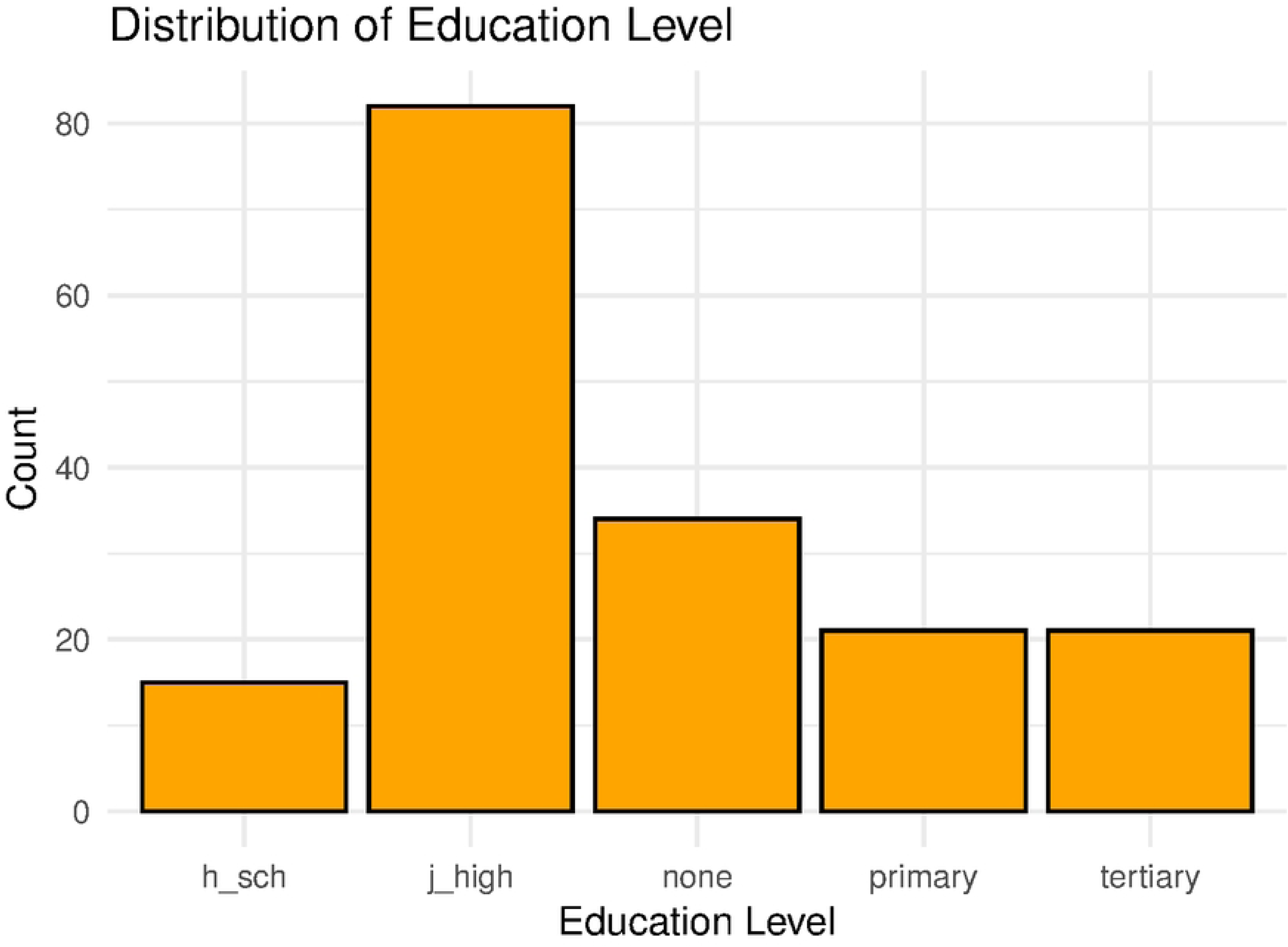
Distribution of educational levels: **Caption**. Frequency count of maternal educational levels to demonstrate highest educational level attained.

**Image_2.**
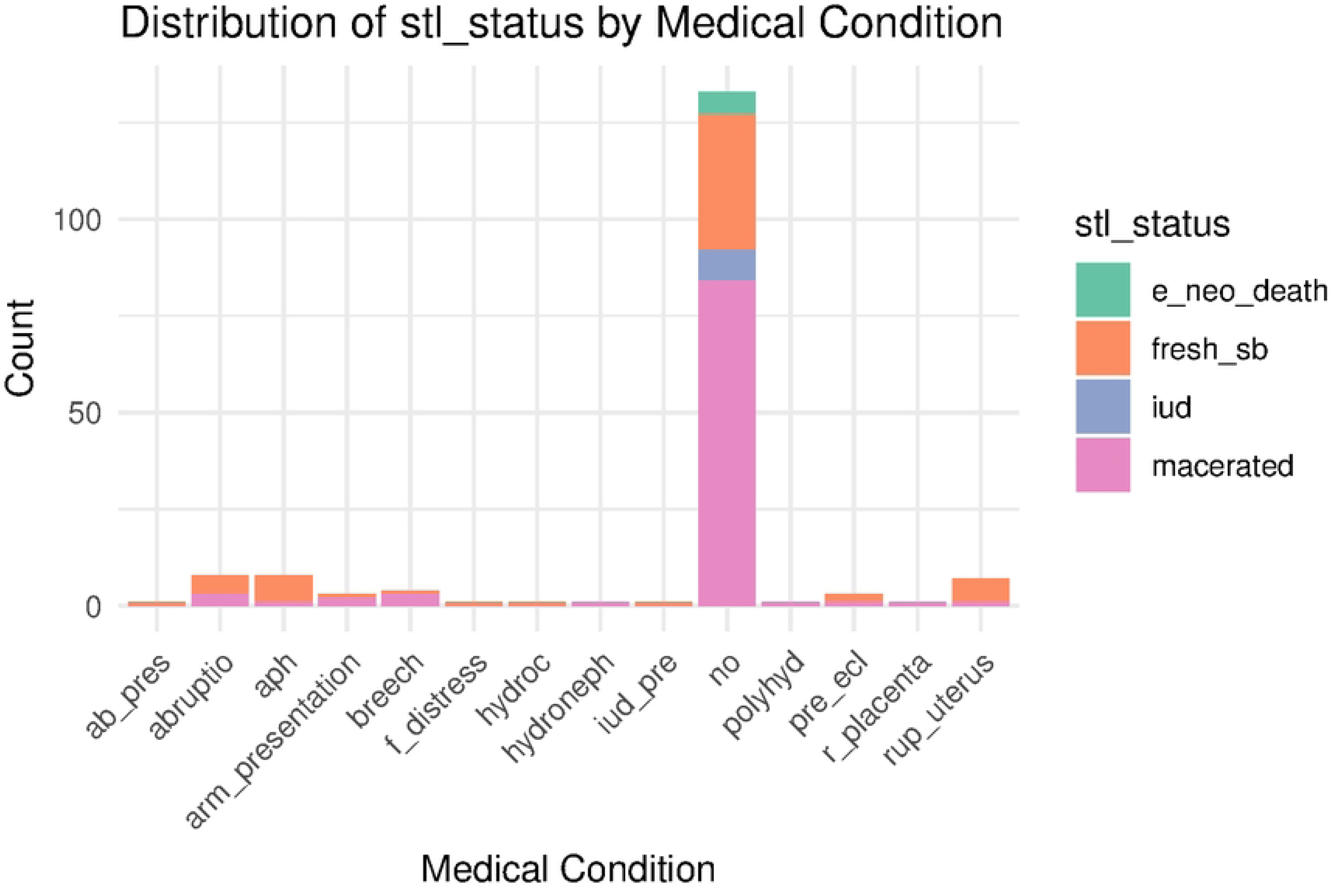
Distribution of stillbirth by medical condition: **Caption.** Describes the various medical conditions recorded for each incidence.

**Image_3.**
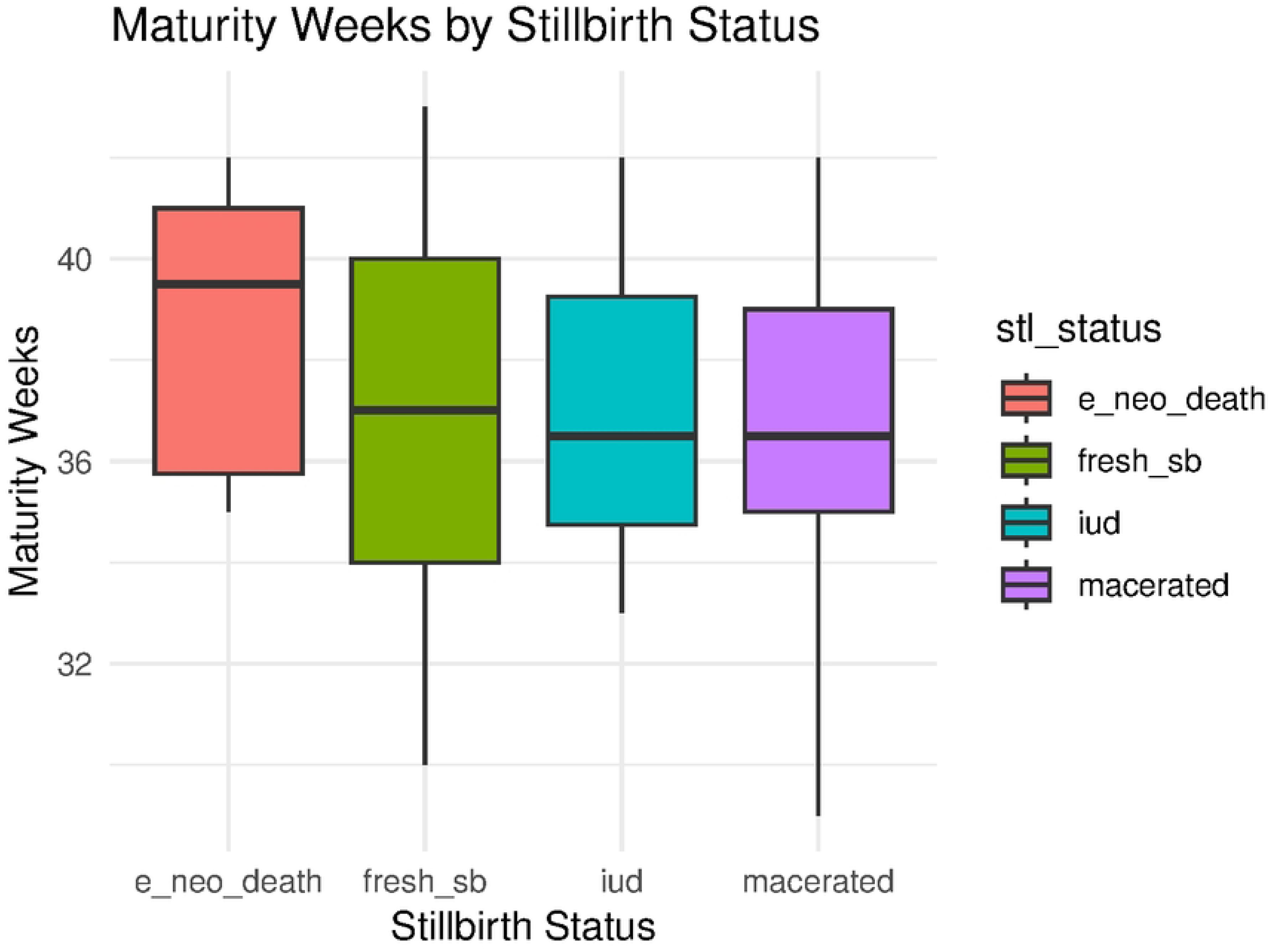
Incidence of stillbirths per maturity: **Caption**. Show incidence of stillbirth per fetal maturity in weeks.

**Image_4.**
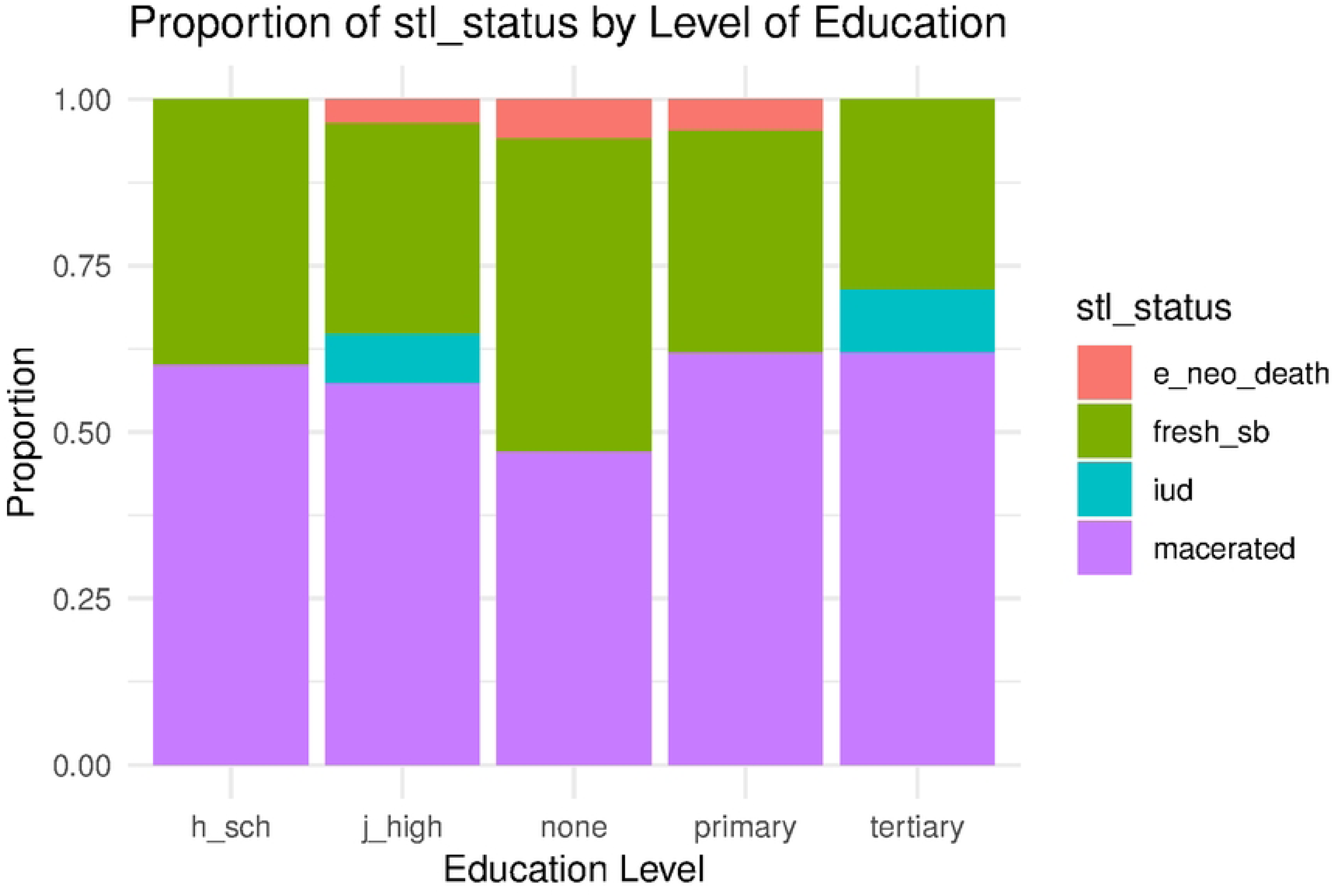
Proportion of stillbirth per educational level: **Caption**. Describe proportion of stillbirths per sampled educational level including those with no formal education.

**Image_5.**
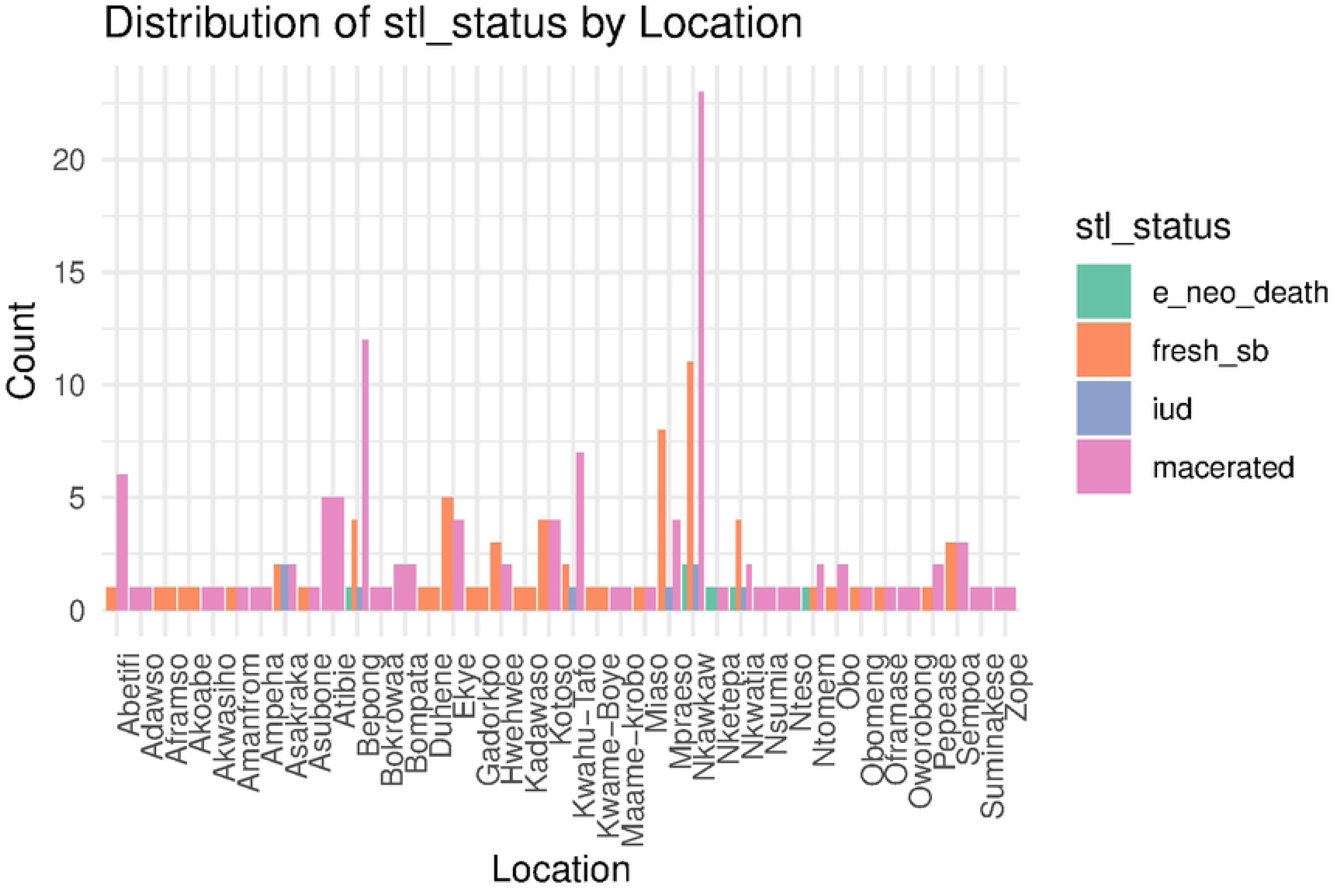
Distribution of stillbirth by geographical area: **Caption**. Distribution of stillbirth per communities sampled.

**Image_6.**
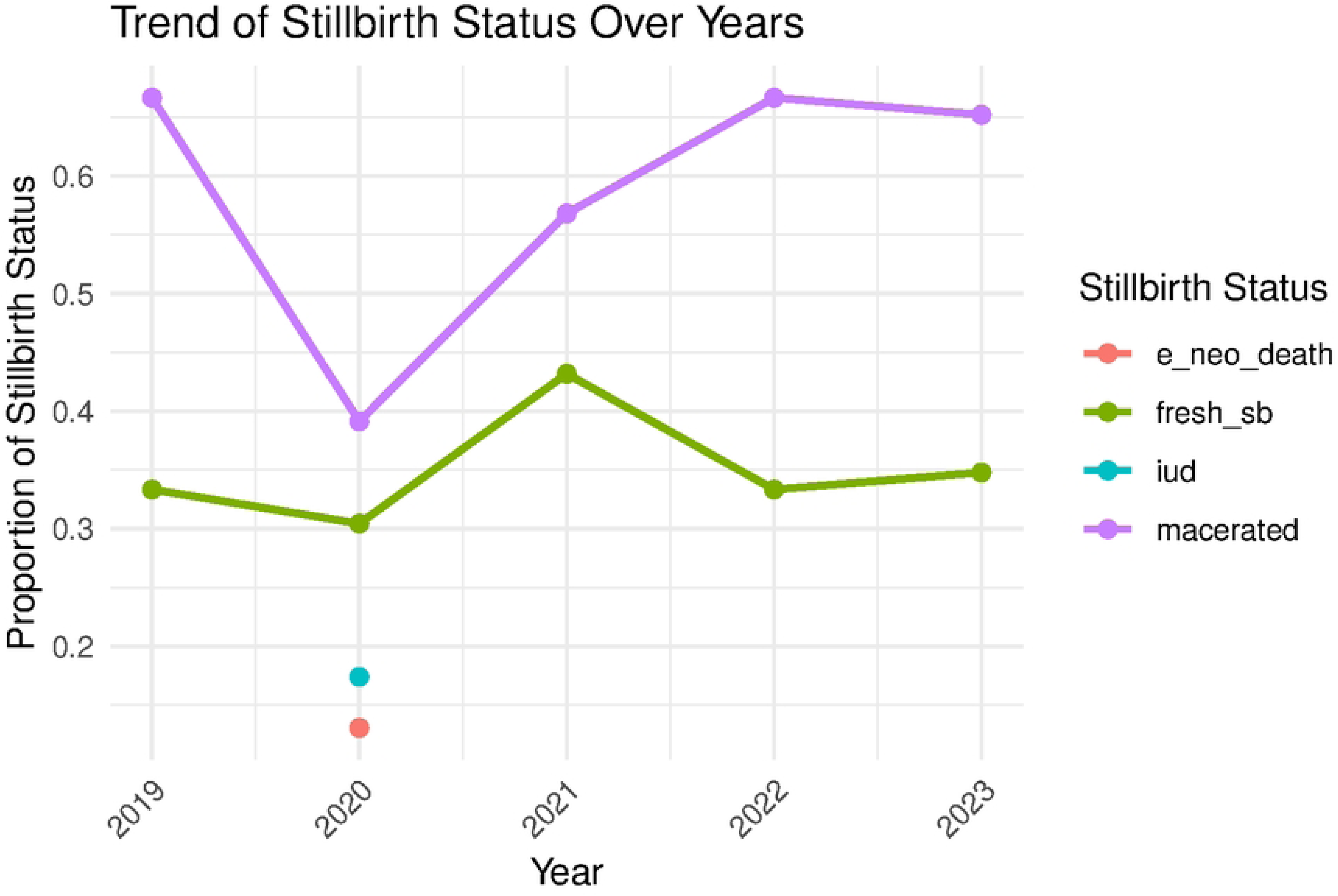
5-year trend analysis: **Caption**. A 5-year trend analysis of incidence of stillbirth, from 2019-2023.

**Image_7.**
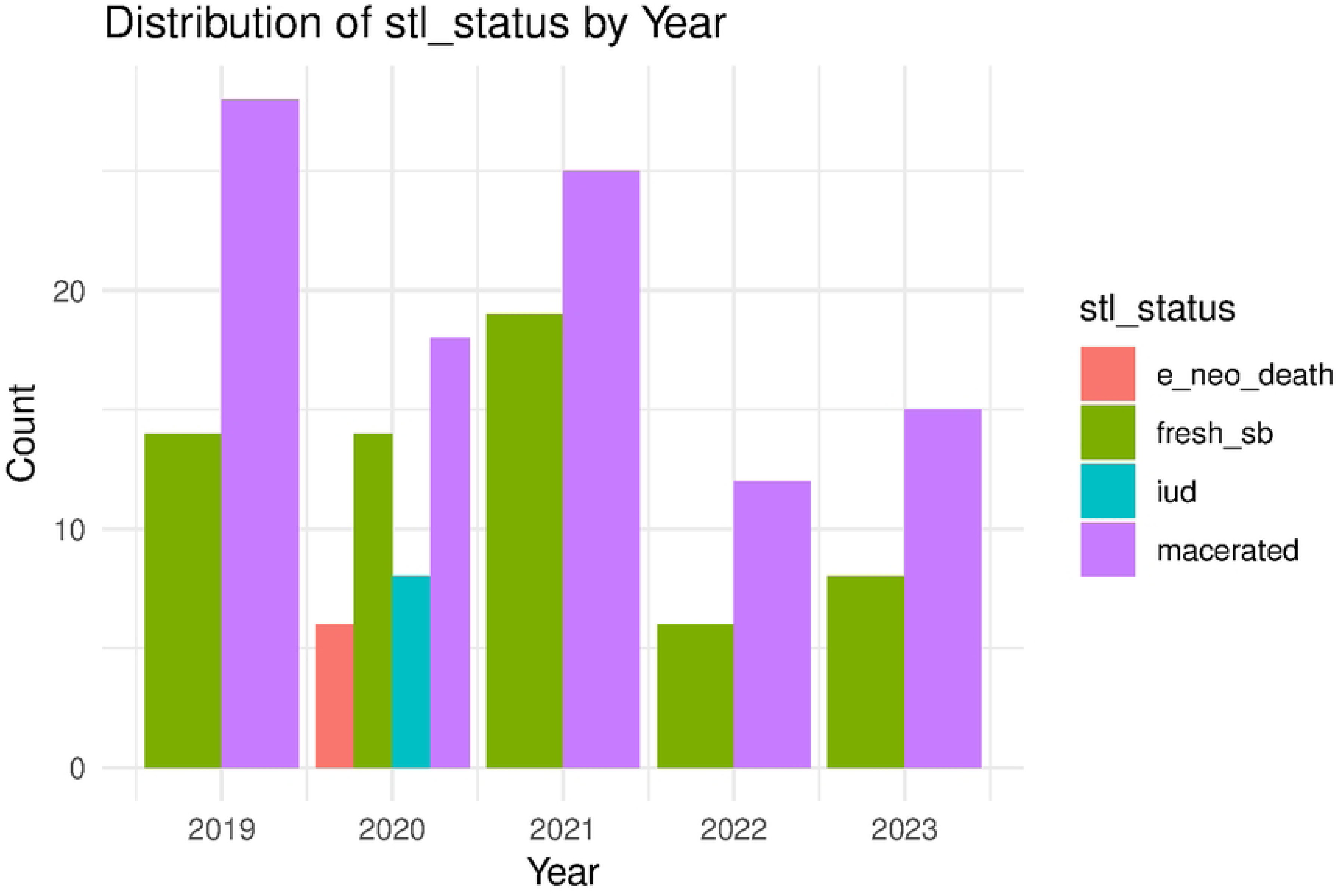
Frequency count of stillbirth over a 5-year period: **Caption**. Frequency distribution of incidence of stillbirths from 2019-2023.

## References

[1] “Stillbirths and stillbirth rates - UNICEF DATA.” https://data.unicef.org/topic/child-survival/stillbirths/ (accessed Dec. 24, 2024).

[2] “Never Forgotten: The situation of stillbirth around the globe - UNICEF DATA.” https://data.unicef.org/resources/never-forgotten-stillbirth-estimates-report/ (accessed Dec. 24, 2024).

[3] UNICEF et al, A neglected tragedy: the global burden of stillbirths - Report of the UN inter-agency Group for Child Mortality Estimation, 2020. 2020. [Online]. Available: https://data.unicef.org/resources/a-neglected-tragedy-stillbirth-estimates-report/

[4] World Health Organization (WHO), “Indicator Sheet Stillbirth Rate”, [Online]. Available: https://www.monitor.srhr.org/related-sheets/Monitor%20Indicator%20sheet%20Stillbirth%20rate.pdf

[5] P. A. Afulani, “Determinants of stillbirths in Ghana: Does quality of antenatal care matter?,” BMC Pregnancy Childbirth, vol. 16, no. 1, pp. 1–17, 2016, doi: 10.1186/s12884-016-0925-9.

[6] V. Gopichandran, S. Subramaniam, and M. J. Kalsingh, “Psycho-social impact of stillbirths on women and their families in Tamil Nadu, India - a qualitative study,” BMC Pregnancy Childbirth, vol. 18, no. 1, pp. 1–13, 2018, doi: 10.1186/s12884-018-1742-0.

[7] P. H. Negandhi et al., “Factors associated with stillbirths in Haryana, India: a qualitative study,” WHO South-East Asia J. public Heal., vol. 7, no. 2, pp. 114–121, 2018, doi: 10.4103/2224-3151.239423.

[8] M. Aminu, “Cause of and Factors Contributing to Stillbirth in sub-Saharan Africa,” no. October, 2017.

[9] A. K. Dah et al., “Stillbirth incidence and determinants in a tertiary health facility in the Volta Region of Ghana,” vol. 3, pp. 1–16, 2023, doi: 10.1371/journal.pone.0296076.

[10] R. Milton et al., “Determinants of Stillbirth From Two Observational Studies Investigating Deliveries in Kano , Nigeria,” vol. 2, no. January, 2022, doi: 10.3389/fgwh.2021.788157.

[11] F. E. Okonofua, L. F. C. Ntoimo, R. Ogu, H. Galadanci, and G. Mohammed, “Prevalence and determinants of stillbirth in Nigerian referral hospitals : a multicentre study,” pp. 1– 9, 2019.

[12] E. Kebede, “Risk factors for stillbirth and early neonatal death : a case-control study in tertiary hospitals in Addis Ababa , Ethiopia,” vol. 8, pp. 1–11, 2021.

[13] M. Bhusal, N. Gautam, A. Lim, and P. Tongkumchum, “Factors associated with stillbirth among pregnant women in nepal,” J. Prev. Med. Public Heal., vol. 52, no. 3, pp. 154–160, 2019, doi: 10.3961/JPMPH.18.270.

[14] S. Ngwenya, B. Jones, D. Mwembe, H. Nare, and A. E. P. Heazell, “The prevalence of and risk factors for stillbirths in women with severe preeclampsia in a high-burden setting at Mpilo Central Hospital , Bulawayo ,” vol. 50, no. 6, pp. 678–683, 2022.

[15] S. P. Malaza, M. S. Maputle, and R. T. Lebese, “Exploring the community perceptions and understanding of stillbirth in South Africa : A qualitative study,” vol. 12, no. 4, pp. 247– 253, 2023.

[16] I. Zile, I. Ebela, and I. Rumba-Rozenfelde, “Maternal risk factors for stillbirth: A registry-based study,” Med., vol. 55, no. 7, pp. 7–13, 2019, doi: 10.3390/medicina55070326.

[17] J. M. Page, N. R. Blue, and R. M. Silver, “Fetal Growth and Stillbirth Stillbirth Fetal death Fetal growth restriction,” vol. 48, p. 8545, 2021.

[18] M. Torkashvand Moradabadi et al., “Investigation of Factors Related to Stillbirth,” Inq. (United States), vol. 61, 2024, doi: 10.1177/00469580241236272.

[19] E. M. Mcclure, S. Saleem, A. Garces, and R. Whitworth, “country study from the Global Network . Let us know how access to this document benefits you,” 2020.

[20] B. Atkins, L. Kindinger, M. P. Mahindra, Z. Moatti, and D. Siassakos, “Stillbirth : prevention and supportive bereavement care,” 2023, doi: 10.1136/bmjmed-2022-000262.

[21] R. Yao, C. V Ananth, B. Y. Park, and L. Pereira, “Obesity and the risk of stillbirth : a population-based cohort study,” Am. J. Obstet. Gynecol., vol. 210, no. 5, pp. 457.e1–457.e9, 2014, doi: 10.1016/j.ajog.2014.01.044.

[22] A. Harnden, R. Mayon-White, D. Mant, D. Kelly, and G. Pearson, “Child deaths: Confidential enquiry into the role and quality of UK primary care,” Br. J. Gen. Pract., vol. 59, no. 568, pp. 819–824, 2009, doi: 10.3399/bjgp09X472520.

[23] E. Malacova et al., “Stillbirth risk prediction using machine learning for a large cohort of births from Western Australia, 1980–2015,” Sci. Rep., vol. 10, no. 1, pp. 1–8, 2020, doi: 10.1038/s41598-020-62210-9.

[24] R. S. Cronin, B. F. Bradford, V. Culling, J. M. D. Thompson, E. A. Mitchell, and L. M. E. McCowan, “Stillbirth research: Recruitment barriers and participant feedback,” Women and Birth, vol. 33, no. 2, pp. 153–160, Mar. 2020, doi: 10.1016/J.WOMBI.2019.03.010.

[25] L. G. Gordon et al., “Healthcare costs of investigations for stillbirth from a population-based study in Australia,” Aust. Heal. Rev., vol. 45, no. 6, pp. 735–744, 2021, doi: 10.1071/AH20291.

[26] S. Mengistu, A. Debella, T. Mulatu, F. Mesfin, K. T. Danusa, and M. Dheresa, “Stillbirth and Associated Factors Among Women Who Gave Birth at Hiwot Fana Specialized University Hospital, Harar, Eastern Ethiopia,” Front. Pediatr., vol. 10, no. May, pp. 1–7, 2022, doi: 10.3389/fped.2022.820308.

[27] “Stillbirth - Causes - NHS.” https://www.nhs.uk/conditions/stillbirth/causes/ (accessed Jan. 04, 2025).

[28] “Talking with Families about Stillbirth | Stillbirth | CDC.” https://www.cdc.gov/stillbirth/hcp/conversation-tips/index.html (accessed Jan. 04, 2025).

[29] M. Lofwander, “Stillbirths and associations with maternal education. A registry study from a regional hospital in north eastern Tanzania,” Epidemiol. Community Health, vol. 66, no. 7, 2012.

[30] G. O. Udigwe, O. A. Onyegbule, I. I. Mbachu, and V. Oguaka, “Prevalence and pattern of stillbirths in a tertiary institution in South-East Nigeria,” Orient J. Med., vol. 25, no. 3–4, pp. 88–93, 2013, [Online]. Available: http://search.ebscohost.com/login.aspx?direct=true&AuthType=cookie,ip,shib&db=awn&AN=ojm-94504&site=ehost-live http://www.ajol.info/index.php/ojm/article/view/94504

[31] N. E. Anyichie and E. N. Nwagu, “Prevalence and maternal socio-demographic factors associated with stillbirth in health facilities in Anambra, south-east Nigeria,” Afr. Health Sci., vol. 19, no. 4, pp. 3055–3062, 2019, doi: 10.4314/ahs.v19i4.27.

[32] O. D. Okochi et al., “Stillbirth at a Nigerian Tertiary Hospital,” Open J. Obstet. Gynecol., vol. 8, no. 8, pp. 756–765, Jul. 2018, doi: 10.4236/OJOG.2018.88079.

[33] J. Muglu et al., “Risks of stillbirth and neonatal death with advancing gestation at term: A systematic review and meta-analysis of cohort studies of 15 million pregnancies,” PLoS Med., vol. 16, no. 7, Jul. 2019, doi: 10.1371/JOURNAL.PMED.1002838.

[34] “Unexplained stillbirth – O&G Magazine.” https://www.ogmagazine.org.au/11/1-11/unexplained-stillbirth/ (accessed Feb. 03, 2025).

[35] C. Bedwell et al., “Understanding the complexities of unexplained stillbirth in sub-Saharan Africa: a mixed-methods study,” *BJOG An Int*. J. Obstet. Gynaecol., vol. 128, no. 7, pp. 1206–1214, 2021, doi: 10.1111/1471-0528.16629.

